# Identification of Genetically Related HCV Infections Among Self-Described Injecting Partnerships

**DOI:** 10.1101/2021.03.22.21254109

**Authors:** Damien C. Tully, Judith A. Hahn, David J. Bean, Jennifer L. Evans, Meghan D. Morris, Kimberly Page, Todd M. Allen

**Affiliations:** Department of Infectious Disease Epidemiology, London School of Hygiene and Tropical Medicine, London, UK; Center for Mathematical Modelling of Infectious Disease, London School of Hygiene and Tropical Medicine, London, UK; Department of Medicine, University of California, San Francisco, California, USA; Ragon Institute of MGH, MIT and Harvard, Cambridge, Massachusetts, USA; Department of Epidemiology and Biostatistics, University of California, San Francisco, California, USA; Department of Internal Medicine, University of New Mexico Health Center, Albuquerque, New Mexico, USA

**Keywords:** Hepatitis C virus, phylogenetic, deep sequencing, injection drug use, molecular epidemiology

## Abstract

**Background:** The current opioid epidemic across the United States has fueled a surge in the rate of new HCV infections among young persons who inject drugs (PWIDs). Paramount to interrupting transmission is targeting these high-risk populations and understanding the underlying network structures facilitating transmission within these communities.

**Methods:** Deep sequencing data were obtained for 52 participants from 32 injecting partnerships enrolled in the UFO Partner Study which is a prospective study of self-described injecting dyad partnerships from a large community-based study of HCV infection in young adult PWIDs from San Francisco. Phylogenetically linked transmission events were identified using traditional genetic-distance measures and viral deep sequence phylogenies reconstructed to determine the statistical support of inferences and the direction of transmission within partnerships.

**Results:** Using deep sequencing data, we found that 12 of 32 partnerships were genetically similar and clustered. Three additional phylogenetic clusters were found describing novel putative transmission links outside of the injecting relationship. Transmission direction was inferred correctly for five partnerships with the incorrect transmission direction inferred in more than 50% of cases. Notably, we observed that phylogenetic linkage was most often associated with a lower number of network partners and involvement in a sexual relationship.

**Conclusions:** Deep sequencing of HCV among self-described injecting partnerships demonstrates that the majority of transmission events originate from outside of the injecting partnership. Furthermore, these findings caution that phylogenetic methods may be unable to routinely infer the direction of transmission among PWIDs especially when transmission events occur in rapid succession within high-risk networks.

**Summary:** Deep sequencing of HCV from 32 self-described injecting partnerships revealed that only 37% were genetically similar and inferring the direction of transmission using phylogenetic tools is challenging as HCV transmission is complex and multifaceted.

## INTRODUCTION

The United States (U.S.) is in the midst of an opioid epidemic which has fueled a surge in HCV incidence, increasing by 294% from 2010-2015 among young persons who inject drugs (PWID) [1–3]. Despite preventive services and curative HCV therapies (direct-acting antivirals, or DAAs), deaths associated with HCV continued to rise above that of 60 other nationally notifiable infectious diseases combined and accounted for almost 20,000 deaths in 2014 [4]. Moreover, national sentinel surveillance of acute HCV incidence rates has detected at least 17 states exceeding the national average with seven states among those with incidence rates twice the national average [2,3]. For example, in Massachusetts in 2011 there was an outbreak of HCV among adolescents injecting prescription opioids and heroin [5] and more recently between 2015 and 2018 Northeastern Massachusetts experienced an outbreak of HIV and HCV attributable to syringe sharing of opioids and homelessness [6,7]. Similarly, in Wisconsin in 2012 a significant increase in statewide HCV infections was associated with increasing injection opioid drug and heroin use and sharing of injection equipment [8]. A more worrying trend is the emerging and rising epidemic among young adult PWID in non-urban areas [9], where an alarming 364% increase in new HCV infections occurred between 2006-2012 among Central Appalachia states [10]. This unprecedented U.S. epidemic has created the impetus for the development of novel public health and treatment intervention strategies to target HCV transmission networks and interrupt transmission. Approaches aimed at using targeted interventions towards members of the community who contribute most and are highly connected to other contacts within the population may be the most efficient way to interrupt HCV dissemination. However, implementation of public health interventions necessitates that the structure of contact and transmission networks is well-defined and that the main drivers of transmission are understood, particularly in the setting of concentrated outbreaks where the conditions that drive outbreaks are often unknown.

In this study, we used a well-defined, sampled cohort of young adult PWIDs from San Francisco to reconstruct the HCV transmission network among self-described injecting partnerships using a deep sequencing approach [11]. We specifically addressed three key questions: 1) we assessed the extent to which the at-risk partners have acquired a genetically similar virus from their self-identified contacts; 2) we evaluated the accuracy of inferring the direction of transmission between linked dyads; and 3) we examined the factors that are associated with the transmission of genetically related infections within the injecting dyads. Together, these analyses serve as an important underpinning for understanding HCV transmission networks at the population level.

## MATERIALS AND METHODS

### Study population and design

Study participants were recruited into the ‘Partner Study’, a sub-study of the UFO study which represents a large prospective community-based epidemiologic study of young adult injectors at risk for HCV in San Francisco, CA [12,13]. From May 2006 to December 2016 UFO study participants were invited to recruit injecting partners to participate in this prospective sub-study of HCV transmission within HCV-serodiscordant injection partnerships [14]. Injection partnerships or ‘dyads’ were eligible for this study if: (i) individuals injected together in the same physical space at least 5 times in the prior month; (ii) they were discordant on HCV RNA or HCV RNA concordant positive with at least one of the partners identified as being acutely infected (HCV RNA positive/anti-HCV negative); and (iii) both members of the dyad had concordant answers to a diverse set of screening questions to validate their injecting activity with their injecting partner. Upon enrollment study participants were asked to return monthly for six-months for follow-up interviews. Re-enrollment for an additional six months occurred if the partnership members were still actively injecting together and remained HCV RNA discordant (meeting the same criteria as above). Partner study participants were allowed to enroll with a maximum of three concurrent injecting partners. The definition of an injecting partnership did not explicitly require that drugs or injecting equipment be shared.

### HCV testing

Anti-HCV antibodies were detected using a third generation EIA (EIA-3; Abbott Laboratories) and HCV RNA testing was performed quarterly using a transcription mediated amplification (TMA) technique (dHCV TMA assay component of the Procleix HIV-1/HCV assay, Gen-Probe Inc., San Diego, CA) to detect early HCV infection in those who tested anti-HCV negative. [13,15].

### Viral RNA extraction and RT-PCR

HCV RNAs were extracted from 140μL of plasma of patient samples following the manufacturers’ protocol for the QIAamp viral RNA mini kit for plasma (Qiagen).

### PCR amplification of Core-NS2, HVR1 and NS5B

For each study participant, a RT-PCR amplification was performed across the Core-NS2 region (H77: 279 - 3542) or the HVR1 (H77: 381 – 1711 (1a), 381 – 1701 (3a)). In addition, a 389-base-pair fragment (H77: 8250 - 8638) of the NS5B region was attempted and amplified from samples. See **Supplementary Methods** for details on primers and PCR conditions used.

### Illumina deep sequencing and data analysis

Purified PCR amplicons were fragmented and barcoded using NexteraXT DNA Library Prep Kit, as per manufacturer’s protocol. Samples were pooled and sequenced on an Illumina MiSeq platform, using a 2 x 250 bp V2 reagent kit. Paired-end reads obtained from Illumina MiSeq were cleaned, *de-novo* assembled and variants called using an in-house bioinformatics pipeline extending from our prior deep sequencing pathogen studies [16,17]. Refer to **Supplementary Methods** for more details on the deep sequencing analysis.

### Phylogenetic reconstruction and cluster analysis

Consensus sequences were aligned using MUSCLE [18] and phylogenetic tress were inferred using maximum likelihood analysis employing the best fit model of nucleotide substitution as implemented within IQ-TREE with 1,000 bootstrap replicates [19]. In order to support the identification of local clusters additional reference sequences from North America were obtained from the HCV-GLUE sequence database (http://hcv-glue.cvr.gla.ac.uk/#/home). Clusters were identified using ClusterPicker v1.2.4 [20] with a bootstrap threshold of 90% and a maximum genetic distance threshold of 0.05 for Core-NS2 and 0.02 for NS5B [21,22].

### Deep sequencing phylogenetic analysis

We utilized phyloscanner (version 1.8.0) [23] to analyze the phylogenetic relationships between and within hosts of all individuals simultaneously using mapped reads produced by Illumina deep sequencing (see **Supplementary Methods** for additional details).

### Ethical approval

All study protocols and procedures were reviewed and approved by the UCSF institutional review board and the institutional review board of Massachusetts General Hospital.

## RESULTS

### Study cohort characteristics

A total of 101 injecting partnerships reflecting 122 unique participants (some at-risk partners were co-enrolled under multiple index cases) were previously enrolled in the UFO partnership study [24] (**Figure. 1)**. Among the 101 injecting partnerships, 40 partnerships (56 participants) demonstrated evidence of incident HCV infection (**Figure. 1**). Of those 56 participants comprising either member of the 40 partnerships in which incident HCV infection was observed, we successfully amplified the Core-NS2 region from 44 subjects (79%) and the NS5B region from 45 subjects (80%) (**Table 1**). For 37 subjects (66%) both regions successfully amplified, and for 52 subjects at least one region amplified. Collectively, from these partnerships in which a new HCV infection was observed we amplified and sequenced data from 32 partnerships. The overall HCV genotype distribution among the 52 individuals were: 1a: 65% (n = 34), 1b: 2% (n = 1), 2a: 2% (n = 1), 2b: 6% (n = 3), 3a: 23% (n = 12) and 4a: 2% (n = 1). Maximum-likelihood phylogenetic trees of the Core-NS2 sequences (**Supplementary Fig. 1a**) and NS5B sequences (**Supplementary Fig. 1b**) illustrate that the viral sequences from injecting partnerships are well-representative of the breadth of genetic diversity observed across hundreds of North American HCV isolates.

**Figure 1.**
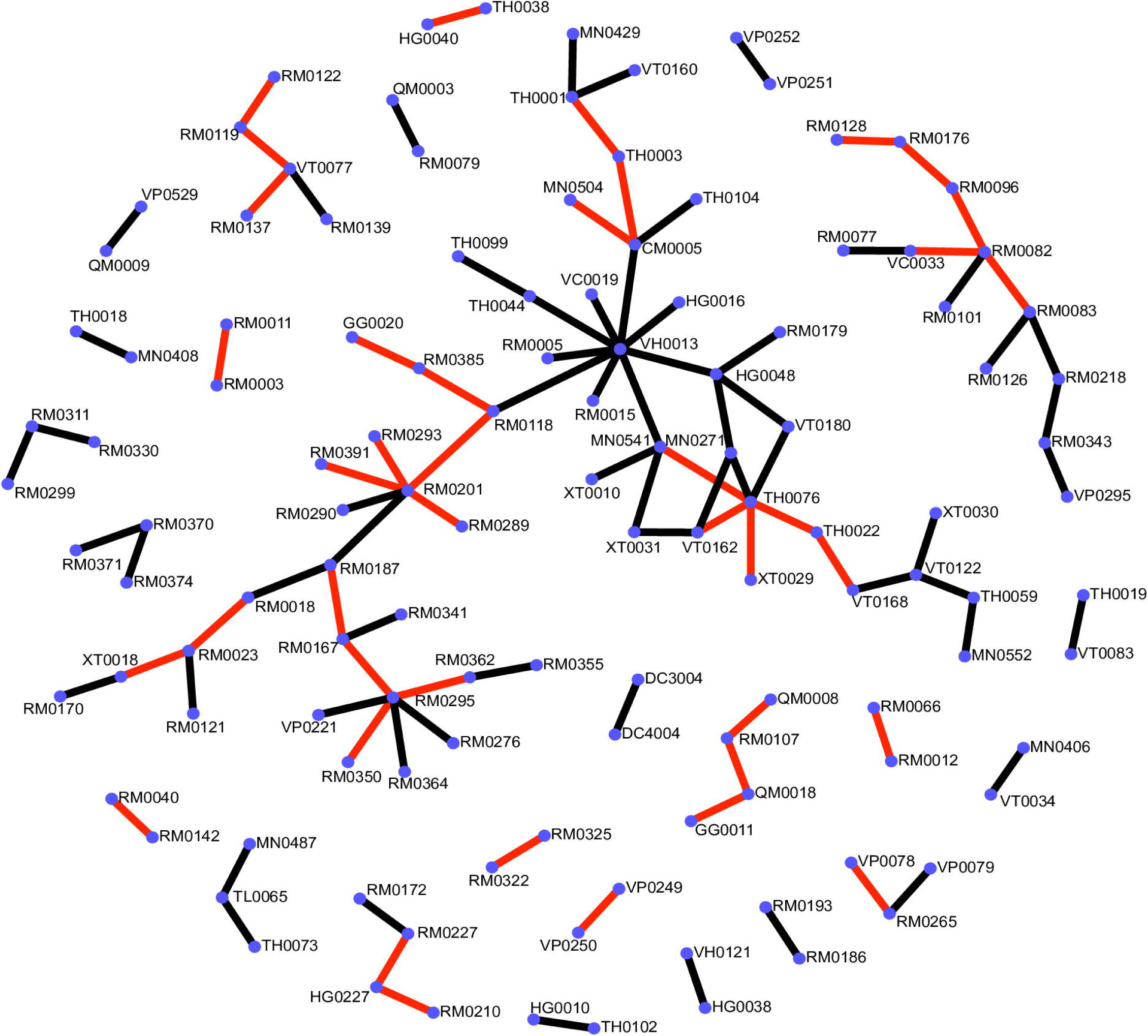
Overview of the study population within the UFO partnership study. 101 partnerships were enrolled and denoted as lines between individuals. Black lines represent those injecting partnerships in which the at-risk partner did not seroconvert. Red lines between study participants reflects those where a new HCV infection was observed in the at-risk partner.

### Phylogenetic clustering using consensus sequences identifies 14 transmission clusters

Phylogenetic analysis of consensus sequences of the Core-NS2 and NS5B regions from the 52 individuals revealed that 52% (27/52) of the participants grouped into 14 clusters. Specifically, phylogenetic analysis of the Core-NS2 region from 44 participants identified 12 clusters (C1-C12) of 25 individuals (**Figure. 2a**). The median genetic distance within the 8 genotype 1a clusters was 0.00468 (IQR: 0.00089 – 0.01) and within the 4 genotype 3a clusters was 0.00016 (IQR: 0 – 0.00056).

**Figure 2.**
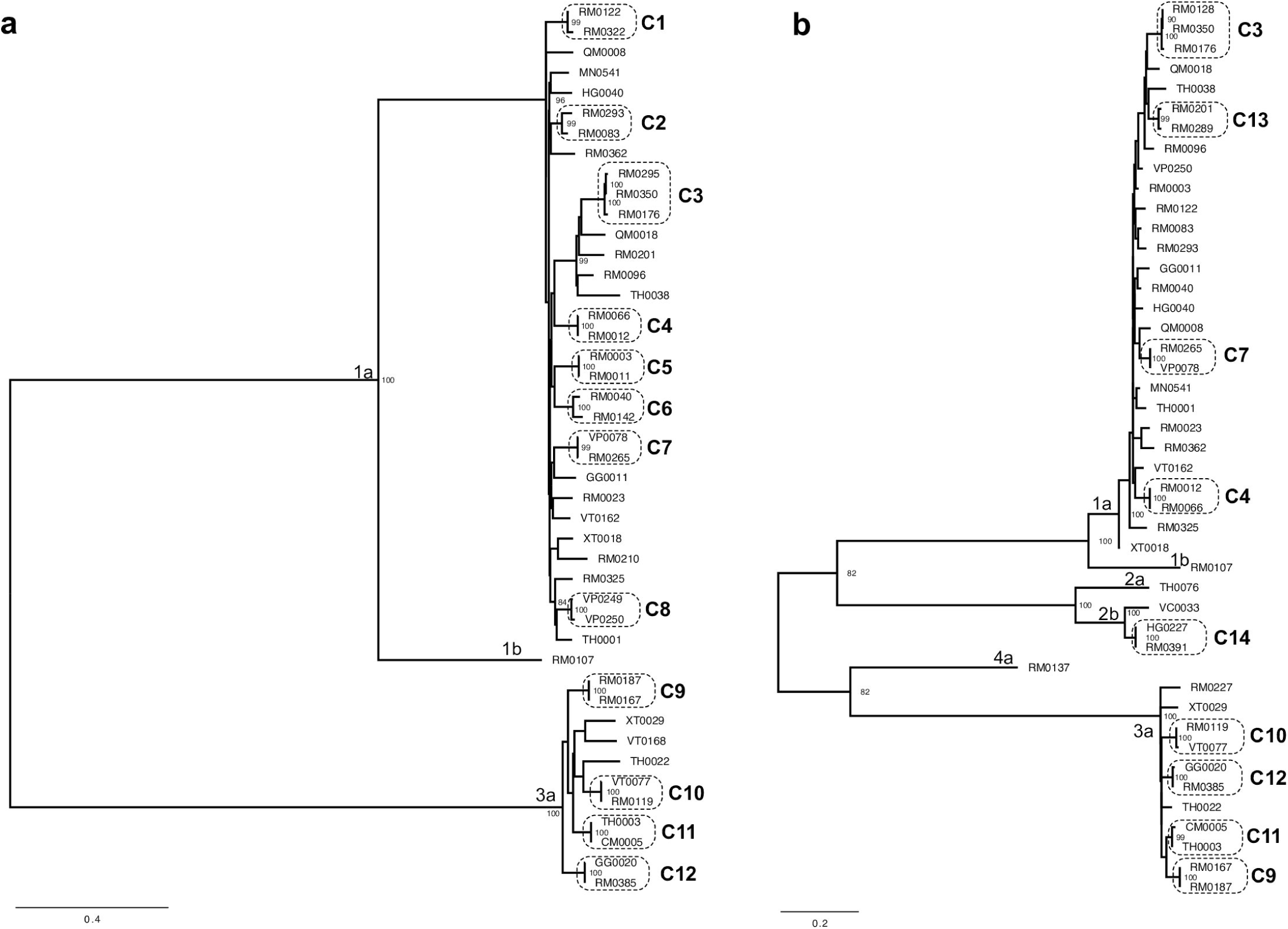
Maximum-Likelihood phylogenetic tree showing phylogenetic clusters within the UFO partnership study for (a) Core-NS2 and (b) NS5B. Phylogenetic clusters defined by bootstrap analysis and genetic distance threshold are highlighted by a dashed line and labelled C1-14. Bootstrap supports values are only shown for nodes over 70%. Genotypes and subtypes are labelled respectively. The scale bar indicates the number of nucleotide substitutions per site.

Phylogenetic analysis of the NS5B region from 45 participants identified 9 clusters at a maximum genetic threshold of 0.02 (**Figure. 2b**), 7 of these were detected in the prior Core-NS2 analysis and 2 were newly found (as Core-NS2 sequence data was not available). The median genetic distance within the 5 genotype 1a clusters was 0.006378 (IQR: 0.001276 – 0.01148). The median genetic distance within the 4 genotype 3 clusters was 0.006378 (IQR: 0.003189 – 0.007653) and for the 1 additional genotype 2 cluster was 0.007653. Cluster 2 was the only cluster that was not found between both sequenced regions due to a low bootstrap support value of 61 within the NS5B region.

### Deep sequencing reveals an additional cryptic genetic linkage within the population

To determine whether incorporation of within-host sequence diversity could improve the resolution of genetically linked clusters we utilized a phylogenetic framework containing both within- and between-host diversity across sliding windows. For each partnership we determined the minimum subgraph distance, defined as the shortest patristic distance between any nodes of one individual within a partnership, across all windows spanning the Core to NS2 region. The distribution of the minimum subgraph distances over the partnerships demonstrated that the majority of index-partner pairs were either phylogenetically closely related (minimum subgraph distance < 0.05 substitutions per site) or distantly related with intermediate distances being rare (**Figure. 3a**). Further inspection of the distribution of subgraph distances found that partnerships could be segregated into those phylogenetically linked and closely related versus phylogenetically unlinked and distantly related (**Figure. 3b**). Partnerships that were phylogenetically close had a median subgraph distance of 0.000001 compared to a median of 0.302 (IQR: 0.109 – 0.881) of those phylogenetically unrelated partnerships. Analysis of the distribution of subgraph distances across NS5B (**Supplementary Figure 2**) revealed a remarkably similar pattern as shown for consensus sequences (**Figure. 2b**) and the Core to NS2 data (**Figure. 3b**).

**Figure 3.**
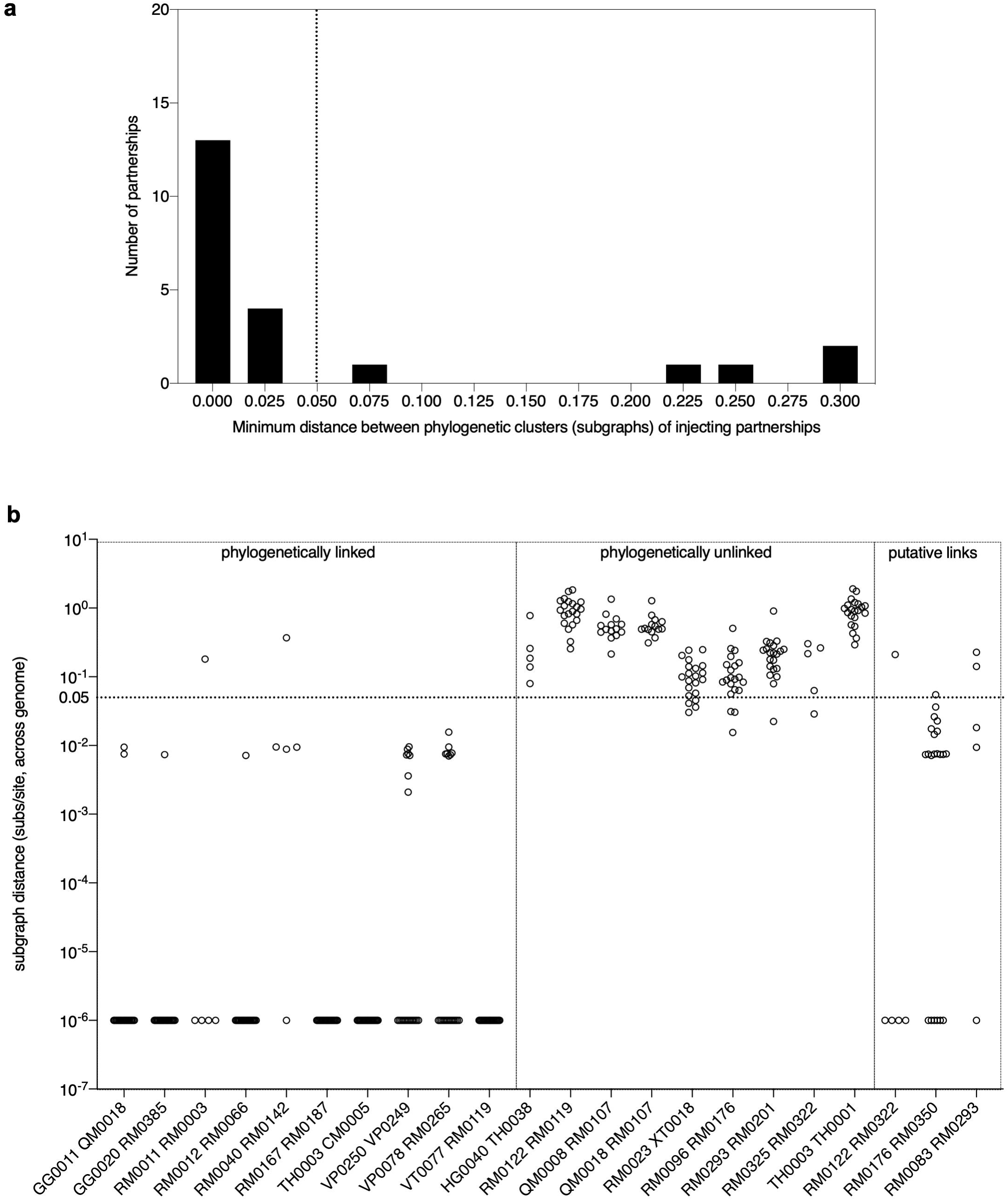
Deep sequence phylogenetic data from injecting partnerships. **(a)** The histogram shows the distribution between injecting partnerships of the minimum subgraph distance obtained using phyloscanner, this analysis included data from 19 self-described partnerships and 3 newly identified putative partnerships. The majority of clustered participants had minimum subgraph distance <0.05 substitutions per site (indicated with a dotted line). **(b)** Subgraph distances calculated from deep sequencing phylogenies for Core to NS2 stratified in those phylogenetically linked and unlinked partnerships. Subgraph distances (y-axis) summarized for all analyzed deep sequence phylogenies for 19 self-described injecting partnerships in which index and at-risk partners have Core to NS2 or HVR sequence data available. Dotted line indicates the distance threshold of 0.05 substitutions per site to define those partnerships classified as phylogenetically close and linked and those phylogenetically distant and unlinked. RM2095 is not depicted as deep sequencing data was not available. Three clusters denoted as putative links as found in figure 2 are also shown.

Together, analyses of the within-host diversity from all deep sequenced participants mirrored our findings using consensus sequences (**Figure. 2**). However, one additional partnership (GG0011 and QM0018), not classified as phylogenetically linked (**Figure. 2**), was found to group in 100% of deep sequence phylogenies where a minor viral variant in QM0018 consistently intermingled with the viral population of GG0011 (**Figure. 3b**). Transmission linkage was independently confirmed within this partnership using the CDC’s global hepatitis outbreak and surveillance technology (GHOST) tool [25] (**Supplementary Fig. 3**).

### Time of sampling between index and partner samples does not impair the ability to detect genetically related infections

We further analyzed whether the time interval between the collection of index and partner samples would have an effect on the ability to detect a genetically related infection. For those phylogenetic linked partnerships, the median time interval between collection of both index and partner samples was 28 days (IQR: 7 – 50.5 days) while for phylogenetic unlinked partnerships the median time interval between collection of samples was 35.5 days (IQR: 16.7 – 255). Thus, we did not observe any significant relationship between duration of sampling between index and partner samples and the inference of phylogenetic linkage (*p = 0*.*301;* **Figure. 4**).

**Figure 4.**
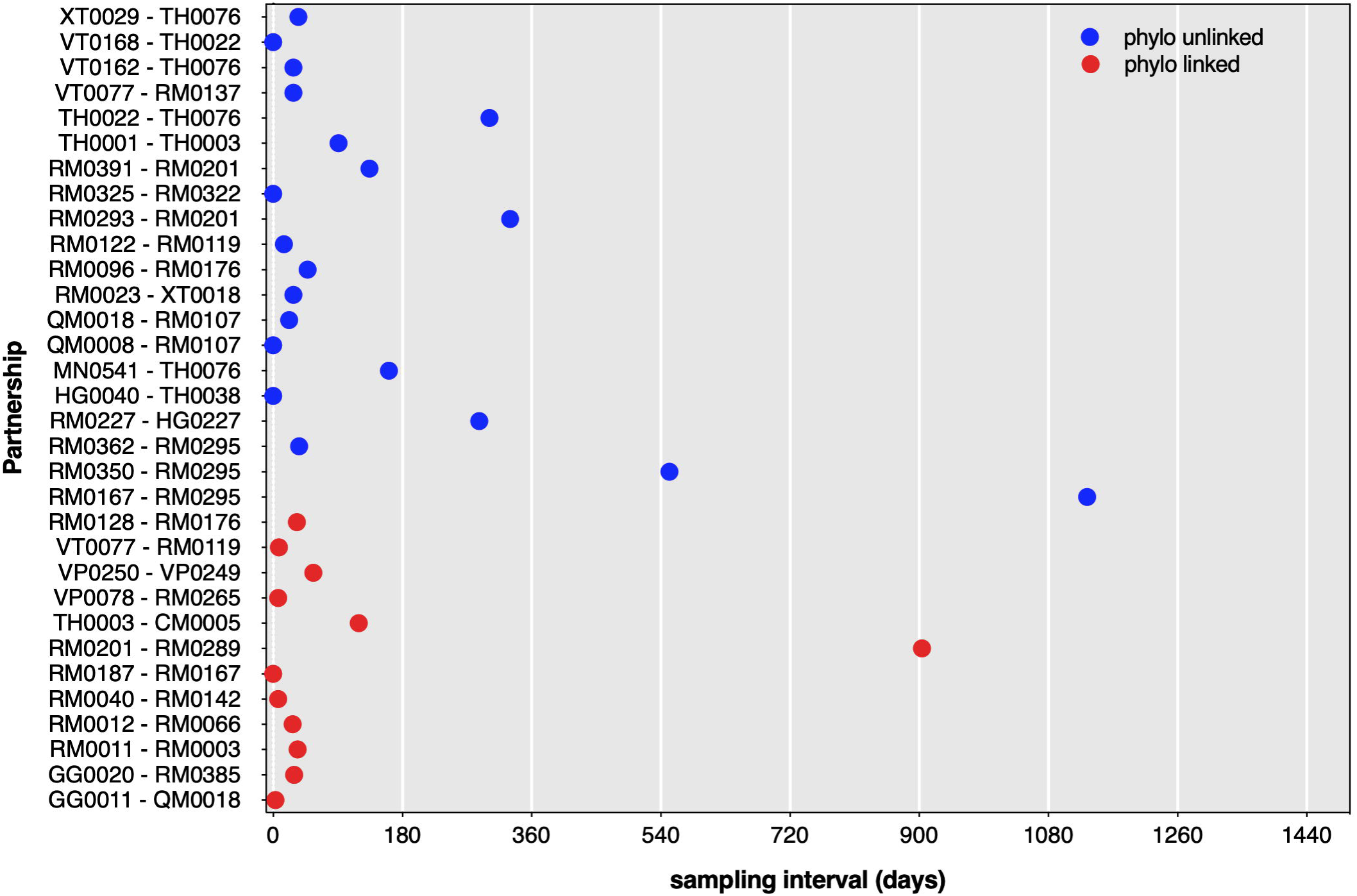
Time between collection of index and partner samples among 32 sequenced self-described partnerships. Blue dots indicate those partnerships that are shown by phylogenetic means to be unlinked; red dots indicate those partnerships that are phylogenetic linked. Data is plotted in days between the collection time of the index and partner samples.

### Inferring transmission direction is challenging among PWIDs due to the close genetic relationship between partnerships

We concentrated on 9 of 12 clustered partnerships in which we had knowledge on the direction of transmission (based on prior negative HCV testing or stage of infection) and evaluated the accuracy of using deep sequencing data to infer transmission direction. Using Core to NS2 sequence data the fraction of pairs with the correct transmission direction (index → partner) was 25% while in 37.5% of partnerships the incorrect direction of transmission was inferred. Three partnerships were classified as linked but no transmission direction could be inferred. In an attempt to increase the accuracy of the inferred transmission direction we examined different window widths and found consistent results (**Supplementary Figure 4)**. At a window width of 310 bp the correct transmission direction was inferred in 38% of partnerships while 38% of partnerships had the incorrect transmission direction inferred. With NS5B data we examined seven partnerships already examined plus the partnership of RM0128 and RM0176 and found that the correct transmission direction was inferred in only one partnership while the remaining sample pairs displayed complex phylogenetic relationships in which tips from each pair are intermingled.

### Onwards transmission of a genetically related infection is supported by dyadic and sexual behavior

To explore whether any factors are associated with the transmission of genetically related infections within our partnerships versus infections originating outside of the partnerships we examined the epidemiological injecting networks between PWIDs (**Figure. 5**). Of the 16 clusters represented, seven are composed of dyads only (i.e. only 1 partner was enrolled by each at-risk person) (**Figure. 5a**) while nine correspond to non-dyads (i.e. >1 partner was enrolled by an at-risk person, or a chain of enrollment occurred, for a total of 3-7 connected persons) (**Figure. 5b**). Of the 32 partnerships in which we have sequence data on both the index and the at-risk partner we confirmed the index and the partner phylogenetically cluster and are genetically similar in 12 partnerships. Three clusters (C1, C2 and C14) represent novel putative links outside of self-described partnerships which share genetic similarity. In addition, one cluster (C3) has expanded to include two other HCV-infected individuals not previously reported to be injecting together (RM0176 from Core to NS2 and RM0350 from NS5B).

**Figure 5.**
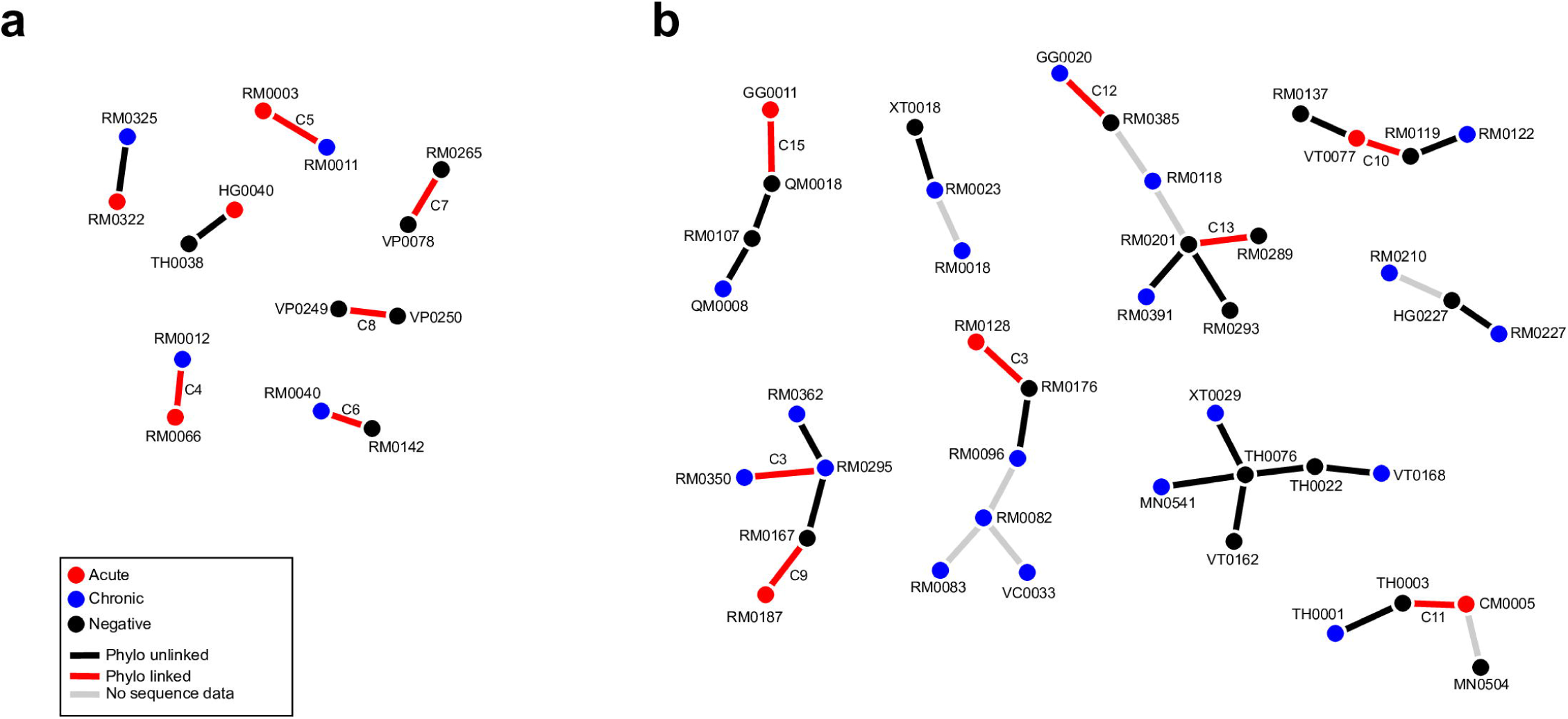
Network representation of the UFO partnerships in which a new HCV infection occurred. Circles and connecting lines denote an injecting partnership in which a new infection occurred. Colored circles denote stage of infection (baseline RNA status) at time of enrollment; blue indicates that the participant was in the chronic stage of infection; red indicates that the participant was in the acute infection window as defined by anti-HCV negative & HCV RNA positive test results; and black indicates that the participant was HCV negative upon study enrollment. Colored lines denote different category membership; red indicates that an injecting partnership was confirmed by sequencing and phylogenetic analysis; black indicates that an injecting partnership was not confirmed by sequencing and phylogenetic analysis; grey lines indicate that an injecting partnership could not be evaluated due to a lack of sequence data for a participant. Phylogenetically defined clusters are labelled as indicated prior and correspond to those as depicted in table 1 and figure 2. **(a)** Dyadic injecting partnerships in which HCV-infected individuals are only enrollment with one at-risk partner **(b)** Larger injecting networks in which infected individuals are linked to multiple index and partners.

Deeper investigation of the phylogenetically linked partnerships revealed that they were predominantly found within those dyads (5 of 7 partnerships (71%)) compared to 5 of 22 (23%) of sample pairs in more complex networks (>2 PWIDs) (**Figure. 6a**). In this cohort phylogenetic linkage within an injecting relationship may be predicated on the size and structure of that relationship, such that dyads are more likely to harbor a virus that is genetically similar compared to those within larger injecting networks (*p = 0*.*03;* OR: 8.5 [1.3 – 57.9]). Moreover, of those 12 injecting partnerships who share a genetically similar virus, 83.3% of were also in a sexual relationship, compared to 21% of those not phylogenetically linked and reported to be in a sexual relationship between each other (*p = 0*.*0008*; OR 19 [3 - 116] **Figure. 6b**). Only three phylogenetically related injecting partnerships (C3: RM0350 – RM0295; C9: RM0167 – RM0187 and C15: QM0018 and GG0011) indicated that they did not engage in sexual behavior relationship at baseline enrollment.

**Figure 6.**
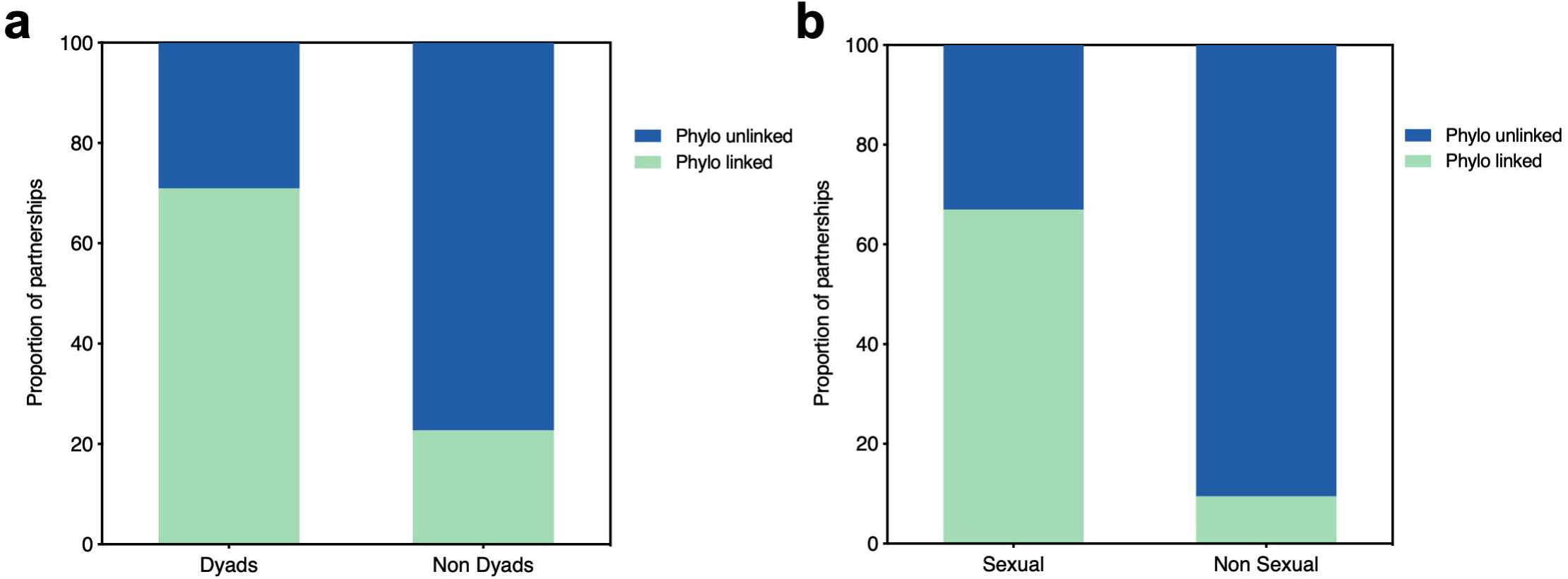
Phylogenetic relatedness of injecting partnerships, cluster size and sexual relationship. **(a)** The proportion of injecting partnerships comparing those PWIDs who are in a dyadic relationship and those that share more than two at-risk partners plotted as a function of their phylogenetic status (linked vs. unlinked). **(b)** The proportion of injecting partnerships comparing whether an index and at-risk partner are engaged in a sexual or non-sexual relationship and their determined phylogenetic status.

## DISCUSSION

In this study we combine high-resolution deep sequencing and phylogenetics with clinical data to investigate the nature of HCV transmission within injecting partnerships. Among these PWIDs we found evidence of clustering suggestive of potential transmission events in only 52% (n = 27) of participants. Within partnerships we found that 63% (n = 20) did not have a genetically related infection. On the other hand, when a partnership was confirmed to be phylogenetically close it was very genetically similar (less than 1% divergence in most cases). This may imply direct transmission or that transmission occurred via unsampled intermediates in quick succession. We found evidence of novel putative transmission links between individuals outside of self-described partnerships and that deep sequencing could enhance the resolution of transmission linkage but not accurately resolve the direction of transmission between infected individuals. Collectively, these findings highlight that the current paradigm of understanding PWID’s risk for infection only within the context of their partnership may be inadequate for understanding the true dynamics of HCV transmission.

The rate of clustering observed in this study was higher than that observed in previous PWID studies [22,26–29] and may be due to the nature of recruiting self-described injecting partnerships, and is more similar to the clustering rate of 54% in Sack-Davis et al who enrolled injecting partnerships [30]. Prior studies have explored the factors associated with phylogenetic clustering and found support for greater clustering with participants of a younger age, HIV co-infection, recent HCV seroconversion, and recent syringe borrowing [22]. Within this study cohort the role of HCV viremia was not directly related to increased transmission but if the index partner was in the HCV-seronegative viremic phase (acute infection) then an increased risk of transmission among partnerships was found suggestive of a complex transmission dynamic among PWIDs [24]. Furthermore, sexual relationships within the UFO cohort have also been associated with increased sharing of syringes and injecting equipment [31] and being in a sexual relationship with one’s partner was associated in an unadjusted statistical analysis with having a phylogenetically linked transmission event [24].

The topological relationship between sequences can potentially be used to infer the direction of transmission [32–34]. To our knowledge this is the first time that such a phylogenetic framework has been applied to HCV. In our case we found that inferring the direction of transmission was more challenging as the virus was heavily intermingled within closely related individuals. In the Core-to NS2 analysis the direction of transmission was inconsistent in 70% of cases with at best only three partnerships demonstrating sufficient evidence for transmission directionality while NS5B only offered with additional partnership that merited support for transmission direction. Thus, among PWIDs the topological signal for direction of transmission may be inherently difficult to disentangle with high confidence as only 4 of 9 pairs (44%) were accurately inferred.

There were several limitations in this study. First, sequence data could not be obtained for all members of the injecting partnership and that our study scope was limited to surveillance in enrolled partnerships in which there was a new HCV infection, and not the wider general community. Second, the measurement of recent sexual behavior represents an indirect representation of intimacy as more detailed partner-specific data such as the timing and frequency of sexual intercourse were not collected. Third, the collection of data via self-reporting can be vulnerable to social desirability bias and may have led to inaccurate or incomplete reporting of partner-specific data. However, self-reported drug and risk behaviors have been shown to be sufficiently reliable [35] but the concordance between specific risk behaviors occurring within injecting dyads may vary [36].

Despite these limitations, our results highlight that HCV transmission in injecting networks is complex and multifaceted with the majority of new infections not being seeded directly from the index case but rather from outside of the reported injection partnership. These results warrant further genomic surveillance among high-risk groups in order to resolve transmission events within networks and guide prevention and treatment modalities. Such necessary steps may help mitigate public health disasters such as that in Scott County, Indiana where HCV was cryptically spreading before the emergence of the large opiate driven outbreak of HIV [37].

## Data Availability

The data that support these findings will be available upon reasonable request.

## FUNDING

This work was supported by the National Institutes of Health [U19 AI082630 and U24 DA044801 to TMA and R01 DA016017 to KP].

## ACKNOWLEDGEMENTS

We thank Matthew Hall for his assistance with Phyloscanner.

## POTENTIAL CONFLICTS OF INTEREST

No authors have any potential conflicts of interests to disclose.

## Supplementary FigureS LEGENDS

**Supplementary Figure 1. Maximum-Likelihood phylogenetic tree of the (a) Core-NS2 region and (b) NS5B region from the North American HCV epidemic**. Phylogenetic tree illustrates that the viral sequences derived from individuals enrolled in the UFO Partner study are well representative of the breadth of genetic diversity circulating across North American HCV epidemic. References sequences are taken from the HCV-GLUE sequence database with sequences belonging to individuals from the UFO cohort labelled as blue circles. Genotypes and subtypes are labelled respectively. The scale indicates substitutions per site and refers to the horizontal branch lengths.

**Supplementary Figure 2. Phyloscanner plot of subgraph distances across bootstrapped phylogenies in NS5B for phylogenetically linked and unlinked partnerships**. Subgraph distances calculated from deep sequencing phylogenies for NS5B stratified into those phylogenetically linked and unlinked partnerships. Subgraph distances (y-axis) summarized for all analyzed deep sequence phylogenies for 12 self-described injecting partnerships in which index and at-risk partners have NS5B amplicon data available. Dotted line indicates the distance threshold of 0.05 substitutions per site to define those partnerships classified as phylogenetically close and linked and those phylogenetically distant and unlinked.

**Supplementary Figure 3. k-step transmission network graph of deep sequencing data from the HVR of QM0018 and GG0011 as outputted by the GHOST web interface**. To be counted as linked by transmission under GHOST the distance has to be smaller than the empirically defined threshold of 3.7%. Each node represents a haplotype with the size of each node proportional to the frequency of the variant it represents, and edge length is proportional to a modified Hamming distance calculation which does not count positions with insertions or deletions as differences. For QM0018 and GG0011 there were 101 and 165 unique variants respectively with 12 variants shared between both samples with a Hamming distance of 0.

**Supplementary Figure 4. Fraction of partnerships with the correct prediction of transmission direction obtained using window widths ranging from 150 bp to 400 bp**. Data is obtained from phyloscanner using deep sequencing data belonging to Core to NS2.

## Notes

### Competing Interest Statement

The authors have declared no competing interest.

## REFERENCES

1. CDC. U.S. 2014 Surveillance Data for Viral Hepatitis | Statistics &amp; Surveillance | Division of Viral Hepatitis | CDC. 2017. Available at: https://www.cdc.gov/hepatitis/statistics/2015surveillance/index.htm#tabs-4-5. Accessed 7 December 2017.

2. Campbell CA, Canary L, Smith N, Teshale E, Ryerson AB, Ward JW. State HCV Incidence and Policies Related to HCV Preventive and Treatment Services for Persons Who Inject Drugs — United States, 2015–2016. MMWR Morb Mortal Wkly Rep 2017; 66:465–469.

3. Zibbell JE, Asher AK, Patel RC, et al. Increases in Acute Hepatitis C Virus Infection Related to a Growing Opioid Epidemic and Associated Injection Drug Use, United States, 2004 to 2014. Am J Public Health 2018; 108:175–181.

4. Ly KN, Hughes EM, Jiles RB, Holmberg SD. Rising Mortality Associated With Hepatitis C Virus in the United States, 2003–2013. Clin Infect Dis 2016; 62:1287–1288.

5. Centers for Disease Control and Prevention (CDC). Hepatitis C Virus Infection Among Adolescents and Young Adults-Massachusetts, 2002-2009. Am J Transplant 2011; 11:1535– 1538.

6. Alpren C, Dawson EL, John B, et al. Opioid Use Fueling HIV Transmission in an Urban Setting: An Outbreak of HIV Infection Among People Who Inject Drugs—Massachusetts, 2015–2018. Am J Public Health 2019; :e1–e8.

7. Cranston K, Alpren C, John B, et al. Notes from the Field: HIV Diagnoses Among Persons Who Inject Drugs - Northeastern Massachusetts, 2015-2018. MMWR Morb Mortal Wkly Rep 2019; 68:253–254.

8. Centers for Disease Control and Prevention (CDC). Notes from the fieldD: hepatitis C virus infections among young adults--rural Wisconsin, 2010. MMWR Morb Mortal Wkly Rep 2012; 61:358.

9. Suryaprasad AG, White JZ, Xu F, et al. Emerging Epidemic of Hepatitis C Virus Infections Among Young Nonurban Persons Who Inject Drugs in the United States, 2006–2012. Clin Infect Dis 2014; 59:1411–1419.

10. Zibbell JE, Iqbal K, Patel RC, et al. Increases in hepatitis C virus infection related to injection drug use among persons aged ≤30 years - Kentucky, Tennessee, Virginia, and West Virginia, 2006-2012. MMWR Morb Mortal Wkly Rep 2015; 64:453–8.

11. Hedegaard DL, Tully DC, Rowe IA, et al. High resolution sequencing of hepatitis C virus reveals limited intra-hepatic compartmentalization in end-stage liver disease. J Hepatol 2017; 66.

12. Hahn JA, PageDShafer K, Lum PJ, et al. Hepatitis C Virus Seroconversion among Young Injection Drug Users: Relationships and Risks. J Infect Dis 2002; 186:1558–1564.

13. Page K, Hahn JA, Evans J, et al. Acute Hepatitis C Virus Infection in Young Adult Injection Drug Users: A Prospective Study of Incident Infection, Resolution, and Reinfection. J Infect Dis 2009; 200:1216–1226.

14. Evans JL, Morris MD, Yu M, Page K, Hahn JA. Concordance of risk behavior reporting within HCV serodiscordant injecting partnerships of young injection drug users in San Francisco, CA. Drug Alcohol Depend 2014; 142:239–44.

15. Page-Shafer K, Pappalardo BL, Tobler LH, et al. Testing strategy to identify cases of acute hepatitis C virus (HCV) infection and to project HCV incidence rates. J Clin Microbiol 2008; 46:499–506.

16. Tully DC, Ogilvie CB, Batorsky RE, et al. Differences in the Selection Bottleneck between Modes of Sexual Transmission Influence the Genetic Composition of the HIV-1 Founder Virus. PLoS Pathog 2016; 12.

17. Henn MR, Boutwell CL, Charlebois P, et al. Whole genome deep sequencing of HIV-1 reveals the impact of early minor variants upon immune recognition during acute infection. PLoS Pathog 2012; 8.

18. Edgar RC. MUSCLE: multiple sequence alignment with high accuracy and high throughput. Nucleic Acids Res 2004; 32:1792–1797.

19. Nguyen L-T, Schmidt HA, von Haeseler A, Minh BQ. IQ-TREE: A Fast and Effective Stochastic Algorithm for Estimating Maximum-Likelihood Phylogenies. Mol Biol Evol 2015; 32:268–274.

20. Ragonnet-Cronin M, Hodcroft E, Hué S, et al. Automated analysis of phylogenetic clusters. BMC Bioinformatics 2013; 14:317.

21. Olmstead AD, Joy JB, Montoya V, et al. A molecular phylogenetics-based approach for identifying recent hepatitis C virus transmission events. Infect Genet Evol 2015; 33:101–109.

22. Jacka, Applegate T, Krajden M, et al. Phylogenetic clustering of hepatitis C virus among people who inject drugs in Vancouver, Canada. Hepatology 2014; 60:1571–1580.

23. Wymant C, Hall M, Ratmann O, et al. PHYLOSCANNER: Inferring Transmission from Within- and Between-Host Pathogen Genetic Diversity. Mol Biol Evol 2018; 35:719–733.

24. Hahn JA, Tully DC, Evans JL, et al. Role of HCV Viremia in Corroborated HCV Transmission Events Within Young Adult Injecting Partnerships. Open forum Infect Dis 2019; 6:ofz125.

25. Longmire AG, Sims S, Rytsareva I, et al. GHOST: global hepatitis outbreak and surveillance technology. BMC Genomics 2017; 18:916.

26. Aitken CK, McCaw RF, Bowden DS, et al. Molecular epidemiology of hepatitis C virus in a social network of injection drug users. J Infect Dis 2004; 190:1586–1595.

27. Bartlett SR, Jacka B, Bull RA, et al. HIV infection and hepatitis C virus genotype 1a are associated with phylogenetic clustering among people with recently acquired hepatitis C virus infection. Infect Genet Evol 2016; 37:252–258.

28. Pilon R, Leonard L, Kim J, et al. Transmission patterns of HIV and hepatitis C virus among networks of people who inject drugs. PLoS One 2011; 6.

29. Hackman J, Falade-Nwulia O, Patel EU, et al. Correlates of hepatitis C viral clustering among people who inject drugs in Baltimore. Infect Genet Evol 2020; 77:104078.

30. Sacks-Davis R, Daraganova G, Aitken C, et al. Hepatitis C Virus Phylogenetic Clustering Is Associated with the Social-Injecting Network in a Cohort of People Who Inject Drugs. PLoS One 2012; 7:e47335.

31. Morris MD, Evans J, Montgomery M, et al. Intimate injection partnerships are at elevated risk of high-risk injecting: a multi-level longitudinal study of HCV-serodiscordant injection partnerships in San Francisco, CA. PLoS One 2014; 9:e109282.

32. Leitner T, Romero-Severson E. Phylogenetic patterns recover known HIV epidemiological relationships and reveal common transmission of multiple variants. Nat Microbiol 2018; 3:983– 988.

33. Ratmann O, Grabowski MK, Hall M, et al. Inferring HIV-1 transmission networks and sources of epidemic spread in Africa with deep-sequence phylogenetic analysis. Nat Commun 2019; 10:1411.

34. Zhang Y, Wymant C, Laeyendecker O, et al. Evaluation of Phylogenetic Methods for Inferring the Direction of Human Immunodeficiency Virus (HIV) Transmission: HIV Prevention Trials Network (HPTN) 052. Clin Infect Dis 2020;

35. Darke S. Self-report among injecting drug users: a review. Drug Alcohol Depend 1998; 51:253–63; discussion 267-8.

36. Evans JL, Morris MD, Yu M, Page K, Hahn JA. Concordance of risk behavior reporting within HCV serodiscordant injecting partnerships of young injection drug users in San Francisco, CA. Drug Alcohol Depend 2014; 142:239–244.

37. Ramachandran S, Thai H, Forbi JC, et al. A large HCV transmission network enabled a fast-growing HIV outbreak in rural Indiana, 2015. EBioMedicine 2018; 37:374–381.

