## Supplementary Methods for "Identification of Genetically Related HCV Infections Among Self-Described Injecting Partnerships"

**PCR amplification of Core-NS2, HVR1 and NS5B.** For Core-NS2, the RT-PCR consisted of 2x First Strand Buffer, sense (177s: CCT TGT GGT ACT GCC TGA TAG) and antisense primers (3542a-1a: GGG YAG CAG TTG ACA CRA TCT or 3542a-3a: CTG GGT AGC CGT AGA AAG CAC CT) at 0.4μM, and Superscript III RT/Platinum *Taq* Mix (Life Technologies), with the following conditions: cDNA synthesis for 30 minutes at 55°C, followed by heat denaturation at 94°C for 2 minutes, the PCR amplification conditions were 40 cycles of 94°C for 15 seconds, 58°C for 30 seconds and 68°C for 4 minutes and a final extension at 68°C for 10 minutes. Amplified products were run on a 1% agarose gel and either PCR purified with the Qiaquck PCR purification kit (Qiagen) or gel extracted and purified using the PureLink quick gel extraction kit (Invitrogen). The HVR1 RT-PCR consisted of 2x First Strand Buffer, 0.4μM of sense and antisense primers, and Superscript III RT/Platinum *Taq* Mix (Life Technologies) with the following conditions: cDNA synthesis for 30 minutes at 55°C, an initial denaturation of 94°C for 2 minutes, 40 cycles of 94°C for 15 seconds, 60°C for 30 seconds, and 68°C for 30 seconds, and a final extension at 68°C for 10 minutes. Amplified products were run on a 1% agarose gel and either PCR purified with the Qiaquick PCR Purification Kit (Qiagen) or gel extracted and purified using the PureLink Quick Gel Extraction Kit (Invitrogen). In addition, a 389-base-pair fragment (H77: 8250 - 8638) of the NS5B region was attempted and amplified from samples using the sense 5′-TTC TCR TAT GAY ACC CGC TGY TTT GA-3′ and reverse 5′-TAC CTV GTC ATA GCC TCC GTG AA-3′ primers. The NS5B RT-PCR consisted of 2x First Strand Buffer, 0.4μM of sense and antisense primers, and Superscript III RT/Platinum *Taq* Mix (Life Technologies) with the following conditions: cDNA synthesis for 30 minutes at 55°C, an initial denaturation of 94°C for 2 minutes, 40 cycles of 94°C for 15 seconds, 56°C for 30 seconds, and 68°C for 60 seconds, and a final extension at 68°C for 10 minutes. Amplified products were run on a 1% agarose gel and either PCR purified with the Qiaquick PCR Purification Kit (Qiagen) or gel extracted and purified using the PureLink Quick Gel Extraction Kit (Invitrogen).

To avoid contamination all PCR procedures were performed under PCR clean room conditions using established procedural protocols including pre-aliquoting of all reagents, use of dedicated equipment and physical separation of sample processing from pre- and post-PCR amplification steps including deep sequencing. Sequencing data was also extensively validated to determine whether contamination was present and phylogenetically examined using phyloscanner.

**GHOST HVR amplification.** To confirm transmission linkage in the GG0011 and QM0018 partnership we sought to employ another deep sequencing approach. For molecular HCV surveillance and outbreak investigation, the CDC have developed a public health tool, Global Hepatitis Outbreak and Surveillance Technology (GHOST), which uses a novel amplicon-based Illumina Miseq sequencing protocol that targets the hypervariable region 1 (HVR1) of the HCV genome. GHOST uses unique haplotypes to measure genetic distance between populations of intra-host HCV HVR1 variants sequenced from each individual and generates a linkage network, where individuals are linked if genetic distance between the HCV populations is below an empirically derived threshold of 3.77%. [1]. Samples were prepared and amplified as previously described. [2]

**Illumina deep sequencing data analysis** In brief, reads were automatically de-multiplexed and duplicate reads were removed using fastuniq v1.1 [3] to limit the influence of PCR artifacts and subsequently quality trimmed using trimmomatic v0.36 [4] if sequencing adapters or low-quality bases (Phred scores < 20) were detected. Vicuna v1.1, a *de novo* consensus assembly algorithm [5], was used to generate consensus assemblies from genetically heterogeneous populations with automated computational finishing and annotation of *de novo* viral assemblies performed using V-FAT v1.1. Lastly, intra-host diversity was inferred with V-Phaser 2 [6].

**Genotyping**. The HCV genotype was determined for all subjects for both Core-NS2 and NS5B sequences using the Oxford HCV automated subtyping tool (http://www.bioafrica.net/rega-genotype/html/subtypinghcv.html).

**Deep sequencing phylogenetic analysis.** Phyloscanner (version 1.8.0) [7] was used to analyze the phylogenetic relationships between and within hosts of all individuals simultaneously using mapped reads produced by Illumina deep sequencing. This method works by incrementally sliding a window along the HCV genome and inferring deep sequence phylogenies for all corresponding reads. The basis of phylogenetic analysis with phyloscanner is the term subgraphs which correspond to tips and internal nodes of a phylogeny assigned to one host that is reconstructed using a maximum-parsimony algorithm. Phyloscanner identifies what within-host subgraphs in these windows and characterizes their distance and topological relationship by aggregating a transmission summary across all windows. Windows were 150 bp and slid across the Core to NS2 region of the HCV genome with a minimum count of 5 reads per individual included in read alignments. This ensures that rare distinct reads were discarded protecting against sequencing error and ensuring that the computational workload was not too expensive. An index and partner sample pair were classified as “linked” if at least 50% of windows demonstrated an ancestral or complex (ancestral relationship but with unknown direction) relationship of any topological type with minimum subgraph distance < 0.05 substitutions per site. A multinomial model as described in Ratmann et al. [8] was used for calculating a score for a link and a score for direction adjusting for windows in which reads from an individual are missing. For the NS5B amplicon we did not split the region into windows to be analyzed separately. Instead, we represented phylogenetic uncertainty by generating a set of 100 phylogenies using RAxML and analyzed these with phyloscanner. The analyses were performed on each phylogeny independently and the results summarized over the entire set. In each phylogeny the shortest patristic distance between subgraphs of reads from two individuals (subgraph distance) was measured to reflect how genetically similar viruses were.

**Direction of transmission inferred from deep sequence data.** Phyloscanner (version 1.8.0) [7] was used to infer the direction of transmission for each partnership. Transmission direction was inferred using windows of different widths (150 to 400 bp in increments of 30 bp with the overlap between windows measured as half of the window width). The inferred transmission direction was considered to be “correct” if index and partner were “linked” and if the correct ancestral relationship (index to partner) was present in ≥37.5% of windows with a minimum subgraph distance of <0.05 substitutions per site. The inferred transmission direction was considered to be “incorrect” if the index and partner were “linked” and if the incorrect ancestral relationships (partner to index) was present in ≥37.5% of windows with a minimum subgraph distance of <0.05 substitutions per site. Injecting partnerships that did not meet either of these criteria were classified as “unknown”. The clinical stage (HCV Naïve: Anti-HCV- and HCV RNA -; Acute HCV: Anti-HCV and HCV RNA +; Chronic HCV: Anti-HCV+ and HCV RNA+ ) of the index and partner from each partnership was examined and used to infer the most probable transmission direction e.g. (1) Index: RNA + , Partner: RNA - ,(2) Index: Chronic HCV, Partner: Acute HCV.

**Supplementary References.**


1. Campo DS, Xia G-L, Dimitrova Z, et al. Accurate Genetic Detection of Hepatitis C Virus Transmissions in Outbreak Settings. J Infect Dis **2016**; 213:957–965.

2. Hochstatter KR, Tully DC, Power KA, et al. Hepatitis C virus transmission clusters in public health and correctional settings, Wisconsin, USA, 2016-2017. Emerg Infect Dis **2021**; 27:480–489.

3. Xu H, Luo X, Qian J, et al. FastUniq: a fast de novo duplicates removal tool for paired

short reads. PLoS One **2012**; 7:e52249.

4. Bolger AM, Lohse M, Usadel B. Trimmomatic: a flexible trimmer for Illumina sequence data. Bioinformatics **2014**; 30:2114–2120.

5. Yang X, Charlebois P, Gnerre S, et al. De novo assembly of highly diverse viral populations. BMC Genomics **2012**; 13:475.

6. Yang X, Charlebois P, Macalalad A, Henn MR, Zody MC. V-Phaser 2: variant inference for viral populations. BMC Genomics **2013**; 14:674.

7. Wymant C, Hall M, Ratmann O, et al. PHYLOSCANNER: Inferring Transmission from Within- and Between-Host Pathogen Genetic Diversity. Mol Biol Evol **2018**; 35:719–733.

8. Ratmann O, Grabowski MK, Hall M, et al. Inferring HIV-1 transmission networks and sources of epidemic spread in Africa with deep-sequence phylogenetic analysis. Nat Commun **2019**; 10:1411.
