## Supplementary figures and images for "Identification of Genetically Related HCV Infections Among Self-Described Injecting Partnerships"

### Supplementary Figure 1

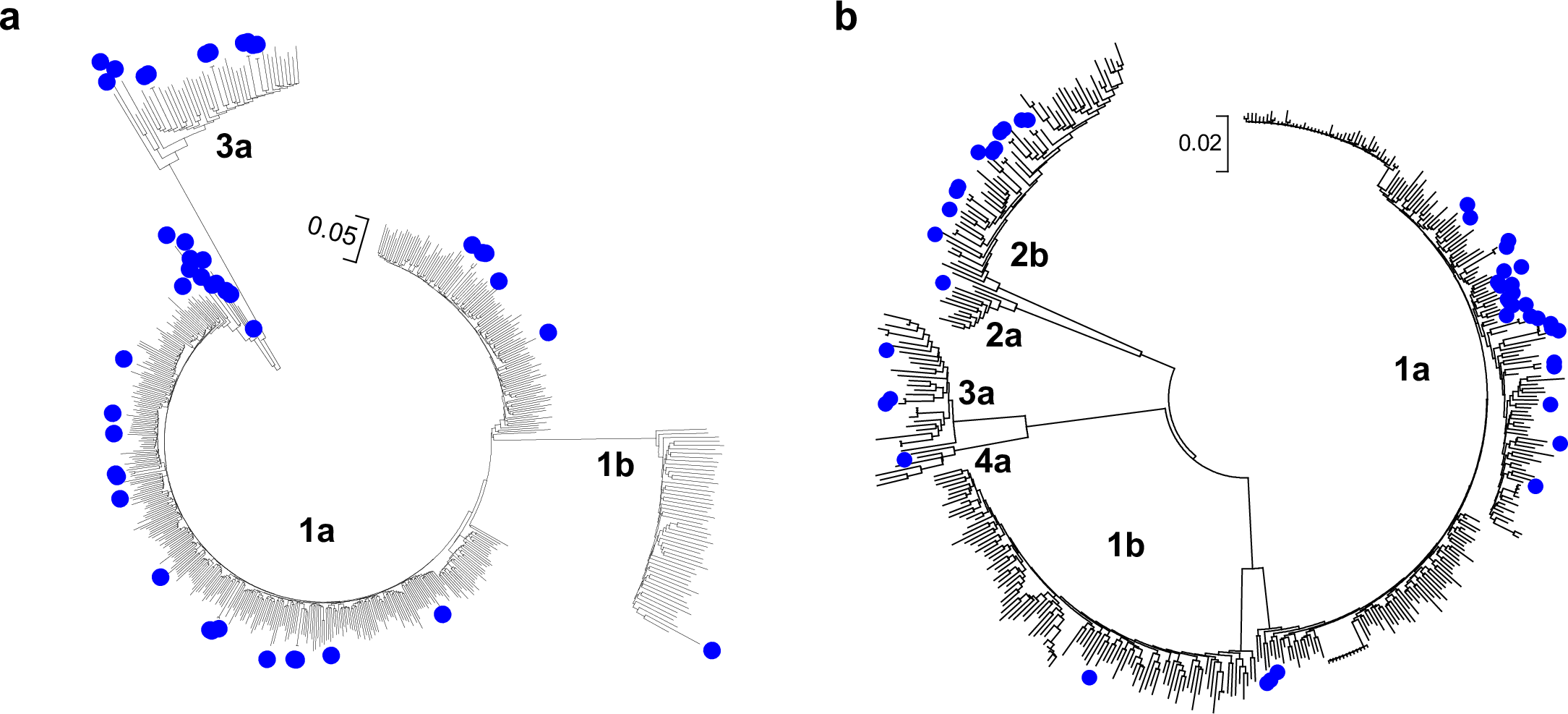

### Supplementary Figure 2

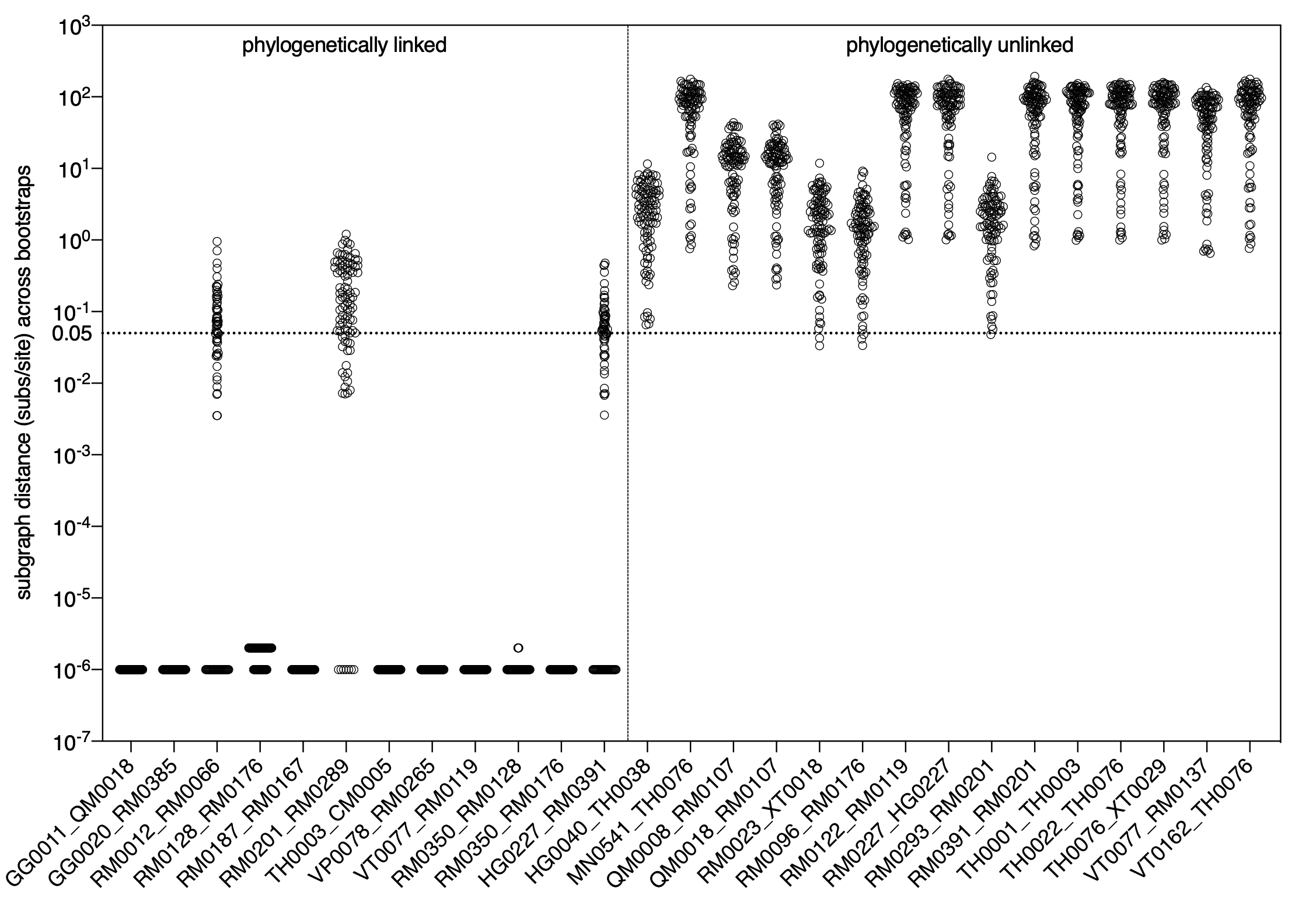

### Supplementary Figure 3

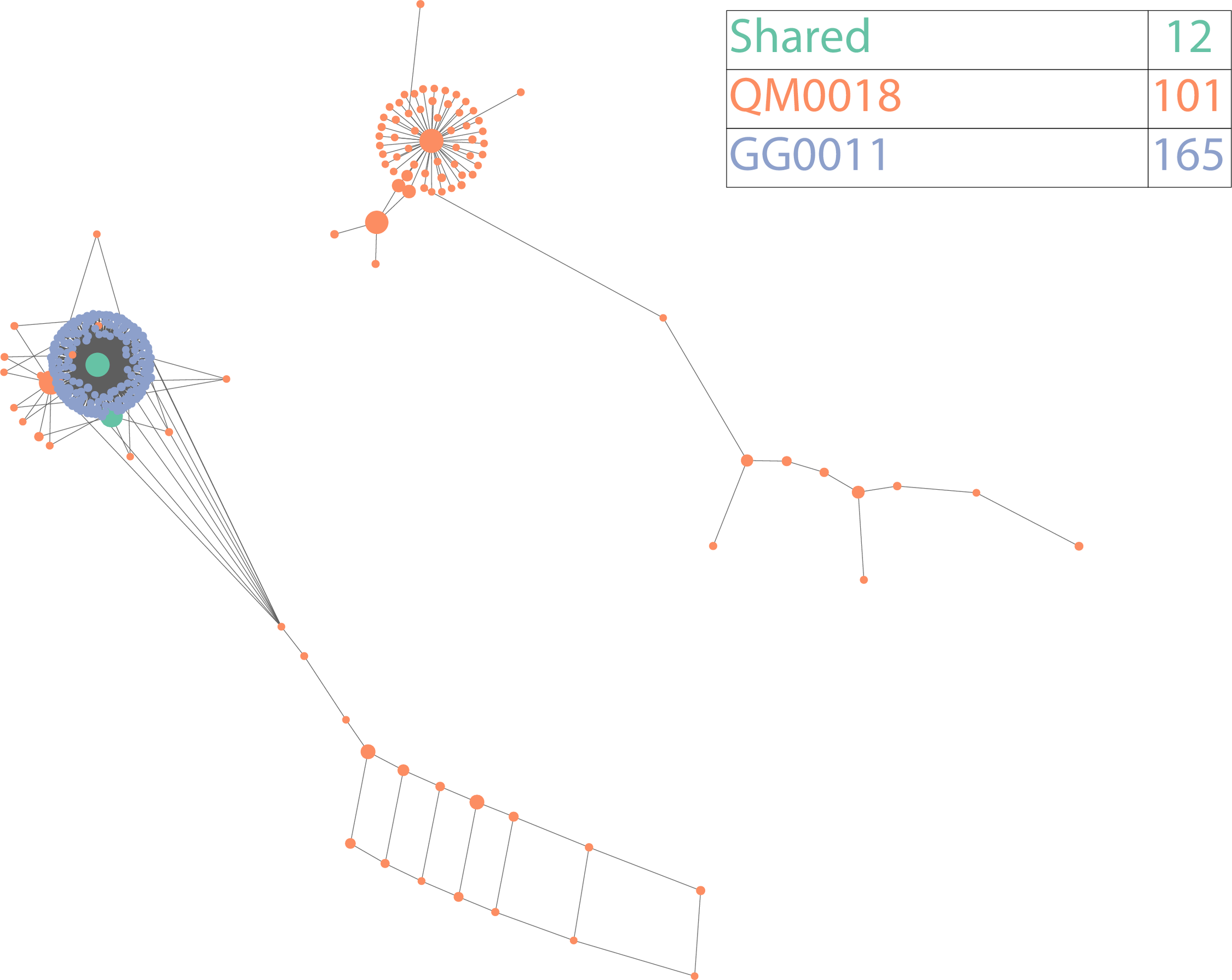

### Supplementary Figure 4

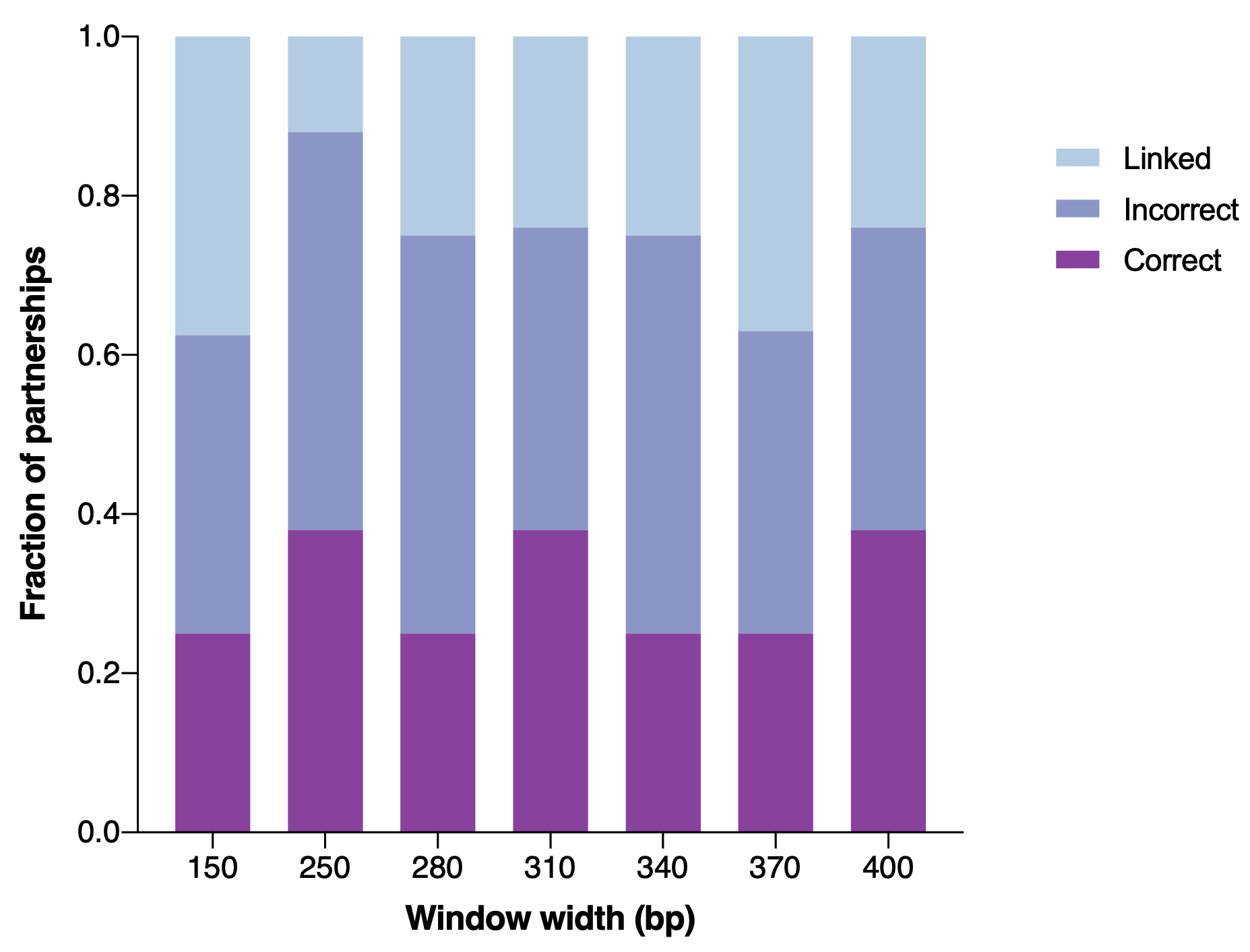
